# Changes in hospital prescribing activity at a specialist children’s hospital during the COVID-19 pandemic - an observational study

**DOI:** 10.1101/2020.12.21.20248153

**Authors:** Emma Vestesson, Carlos Alonso, John Booth, Neil J Sebire, Adam Steventon, Steve Tomlin, Joseph F Standing

## Abstract

**Objective:** To compare hospital activity, patient casemix and medication prescribing and administration before and during the COVID-19 emergency.

**Design:** Retrospective observational study

**Setting:** A specialist children’s hospital in the UK

**Patients:** Inpatients aged 25 and younger treated at a specialist children’s hospital between 29 April 2019 and 6 September 2020

**Results:** There were 21,471 day cases and inpatients treated during the 16 month study period. Day cases (no overnight stay) dropped by around 37% per week. Both admissions and discharges for inpatients (at least one overnight stay) decreased leading to a small reduction in hospital bed days but no reduction in hospital bed nights. The effect on hospital activity on different patient groups varied substantially with some groups such as medical oncology day cases increasing by 13%. As a result, the patient case mix in the hospital was very different during the pandemic. Overall weekly medication administrations decreased for day cases and inpatients, but weekly medication administrations per bed day increased by 10% for day cases and 6% for inpatients.

**Conclusions:** Despite not being badly affected by the disease itself, specialist paediatric hospital services have been greatly affected by the pandemic. The average number of medications per inpatient bed day increased, likely reflecting changes to the patient population, with only those children with severe conditions being treated during the pandemic. These data demonstrate the complex pattern of implications on specialist services and provide evidence for planning the impact of future emergencies and resolution strategies.

## Introduction

The COVID-19 emergency has had wide ranging effects on access to health care in the UK. Despite <1% of COVID-19 cases appearing in children in the first wave and COVID-19 in children being associated with a milder disease progression, most children were still affected by wide-ranging changes to society [3]. A nationwide lockdown where everyone but key workers was recommended to stay at home was implemented on 23 March 2020 and schools had been closed a few days earlier, on 20 March 2020. NHS hospitals in England were instructed to suspend all non-urgent elective surgery by 15 April 2020.

As of April 2020, the majority of complex paediatric inpatients in North Central London CCG were being cared for at Great Ormond Street Hospital (GOSH), a paediatric tertiary care hospital based in central London [4]. This was part of a systems response by north and central London health and care providers to the COVID-19 pandemic aiming to optimize capacity for adults with COVID-19 in other hospitals. Patients at GOSH generally have complex underlying medical conditions with around three quarters of inpatients registered with the hospital being considered as ‘vulnerable’ to COVID-19 according to COVID-19 guidance [5]. There were also changes to working patterns at GOSH with some staff working remotely, more staff being off sick or being transferred to other hospitals.

A large district general hospital in Manchester, UK, reported a 30% decrease in visits to the paediatric emergency department compared to pre-pandemic [6], and a survey of paediatricians in the UK and Ireland reported that over 30% saw patients in emergency departments or paediatric assessment units presenting later than they would have expected prior to the COVID-19 emergency[7]. However, a multi-centre study from the UK examining presentations to paediatric emergency departments at seven hospitals found that the majority (>90%) of presentations that did occur were not delayed[8].

Similar patterns have been reported internationally, such as a 47.2% decrease in presentations to emergency departments in Victoria, Australia [9] and 70-90% reduction in attendances to five paediatric emergency departments in Italy compared to previous data [10].

Though there are studies emerging that look at access and utilisation of emergency health care for children during the COVID-19 emergency, there is a paucity on the impact of changes to care in hospital for paediatric inpatients. Health care utilisation in these different health care settings will be driven by various factors, especially at GOSH where there is no A&E department.

Understanding how inpatient care at a specialist paediatric hospital changed during the pandemic will be important in interpreting the possible drivers for changes in healthcare utilisation such as prescribing patterns during that period.

Using routinely collected, non-identifiable data derived from the electronic health record at GOSH, we conducted a retrospective study to quantify how the COVID-19 emergency affected a specialist children’s hospital. The aim was to compare hospital activity, types of activity, patient case mix and in particular medication prescribing and administration before and during the COVID-19 emergency.

## Methods

### Data sources and study populations

We retrospectively used routinely collected deidentified hospital data (REC approval 17/LO/0008) from children who were inpatients at GOSH between Monday 29 April 2019 and Sunday 6 September 2020. A new electronic health record system went live at GOSH in mid-April 2019, making Monday 29 April 2019 the first full week with data from the new system. The end of the study period coincides with schools in the UK re-opening. Admissions data with information on admission and discharge dates was linked to data on treatment speciality, surgical encounters and medication prescribing data using three variables: a pseudonomised patient ID and the date and time of admission and discharge. Patients older than 25 years of age when admitted were excluded from the study (<1% of admissions) but no other exclusion criteria were applied to the data.

### Outcome measures

Descriptive statistics of patient characteristics recorded at admission, type and source of admission, number of surgical encounters and medication administrations were calculated before and after the start of the COVID-19 emergency based on admission dates.

Patients were included in the COVID-19 pandemic period based on admission dates and some patients will have been admitted before 23 March 2020 but discharged after. These patients were included in the pre-COVID-19 pandemic group.

All data relating to drug administrations were retrieved. The total number of drugs administered was calculated at hospital level. Drug administration per patient bed day was calculated for the most common therapeutic classes.

Patient bed days by specialised service were calculated for specialities with at least 1000 admissions in the study period. The treatment function code was used to identify within which specialised service the patients was admitted to [11].

Length of stay was calculated as the difference in days between admission and discharge. Patients that were not discharged during the study period were censored on the 05 September 2020 and this date was used to calculate length of stay.

Censored patients were not included in the count of day cases or discharges but were included in admission counts, bed days and bed nights.

### Hospital level measures

Weekly counts of admissions, discharges, hospital bed nights, surgical encounters, and medication administrations were used to calculate summary statistics comparing hospital level activity before and during the COVID-19 emergency. All weekly measures were calculated from Monday to Sunday with Monday 23 March 2020 marking the start of the COVID-19 emergency.

Seven day rolling averages were calculated for day cases, inpatient admissions and inpatient discharges.

There were 48 weeks included in the pre-COVID-19 period and 24 in the COVID-19 period.

### Statistical methods

All measures were reported separately for day cases, (defined as patients who were admitted but did not stay overnight), and inpatients (defined as patients admitted and with at least on overnight stay).

Wilcoxon rank-sum tests and chi-square test of independence were used to test for statistically significant differences between the two groups, before and during the COVID-19 emergency, at the 95% confidence level.

All analyses were performed using R 4.0.2 [12].

## Results

The study included a total of 21,471 patients with 52,256 admissions, 1,734,794 medication administrations and 18,719 surgical encounters. For day cases, there were 14,464 individuals, 35,817 admissions, 86,502 medication administrations and 6,009 surgical encounters. For inpatients, there were 10,914 individuals, 16,439 admissions, 1,648,292 medication administrations and 12,710 surgical encounters.

There were 35,817 day cases (14,464 individual patients) and 16,439 inpatient admissions (10,914 individual patients). To compare changes to hospital level activity, median weekly counts of measures of interest before and during COVID-19 were calculated (Table 1). The median number of day cases per week decreased from 588 (IQR 557-618) before to 370 (IQR 280-418) during the COVID-19 emergency (Wilcoxon rank-sum test p<0.001). Medication administrations and surgical encounters also decreased significantly during the COVID-19 pandemic, but weekly medication administrations per bed day increased whilst surgical encounters per bed day was almost halved (Table 1).

**Table 1:**
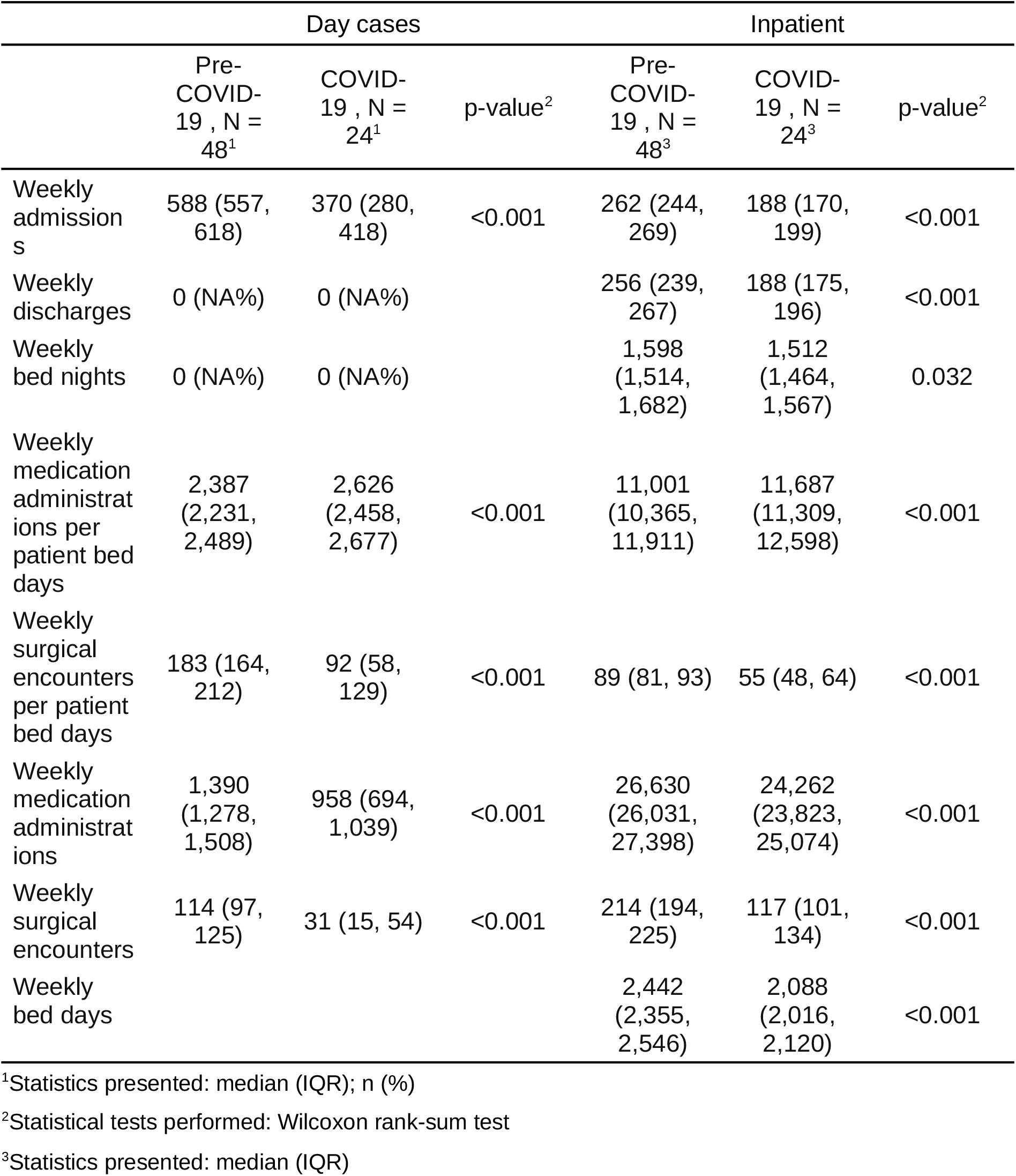
Weekly hospital level activity before and after the start of the COVID-19 emergency

|  | Day cases |  |  | Inpatient |  |  |
| --- | --- | --- | --- | --- | --- | --- |
|  | Pre-COVID-19 , N = 48<sup>1</sup> | COVID-19 , N = 24<sup>1</sup> | p-value<sup>2</sup> | Pre-COVID-19 , N = 48<sup>3</sup> | COVID-19 , N = 24<sup>3</sup> | p-value<sup>2</sup> |
| Weekly admissions | 588 (557, 618) | 370 (280, 418) | <0.001 | 262 (244, 269) | 188 (170, 199) | <0.001 |
| Weekly discharges | 0 (NA%) | 0 (NA%) |  | 256 (239, 267) | 188 (175, 196) | <0.001 |
| Weekly bed nights | 0 (NA%) | 0 (NA%) |  | 1,598 (1,514, 1,682) | 1,512 (1,464, 1,567) | 0.032 |
| Weekly medication administrations per patient bed days | 2,387 (2,231, 2,489) | 2,626 (2,458, 2,677) | <0.001 | 11,001 (10,365, 11,911) | 11,687 (11,309, 12,598) | <0.001 |
| Weekly surgical encounters per patient bed days | 183 (164, 212) | 92 (58, 129) | <0.001 | 89 (81, 93) | 55 (48, 64) | <0.001 |
| Weekly medication administrations | 1,390 (1,278, 1,508) | 958 (694, 1,039) | <0.001 | 26,630 (26,031, 27,398) | 24,262 (23,823, 25,074) | <0.001 |
| Weekly surgical encounters | 114 (97, 125) | 31 (15, 54) | <0.001 | 214 (194, 225) | 117 (101, 134) | <0.001 |
| Weekly bed days |  |  |  | 2,442 (2,355, 2,546) | 2,088 (2,016, 2,120) | <0.001 |

<sup>1</sup>Statistics presented: median (IQR); n (%)
<sup>2</sup>Statistical tests performed: Wilcoxon rank-sum test
<sup>3</sup>Statistics presented: median (IQR)

For inpatients, average weekly admissions and discharges both decreased, whereas there was no statistically significant difference in the median number of bed nights per week. Medication administrations decreased from 26,630 (IQR 26,031-27,398) per week before the pandemic to 24,262 (IQR 23,823-25,074) during, (Wilcoxon rank-sum test p<0.001) but weekly medication administrations per bed day increased.

There was a sharp decrease in the number of day cases in March followed by a slow increase (Figure 1a). Inpatients admissions and discharges show similar patterns, both dropping in March 2020 with gradual recovery. Occupancy levels remained stable during the study period (Figure 1b).

**Figure 1:**
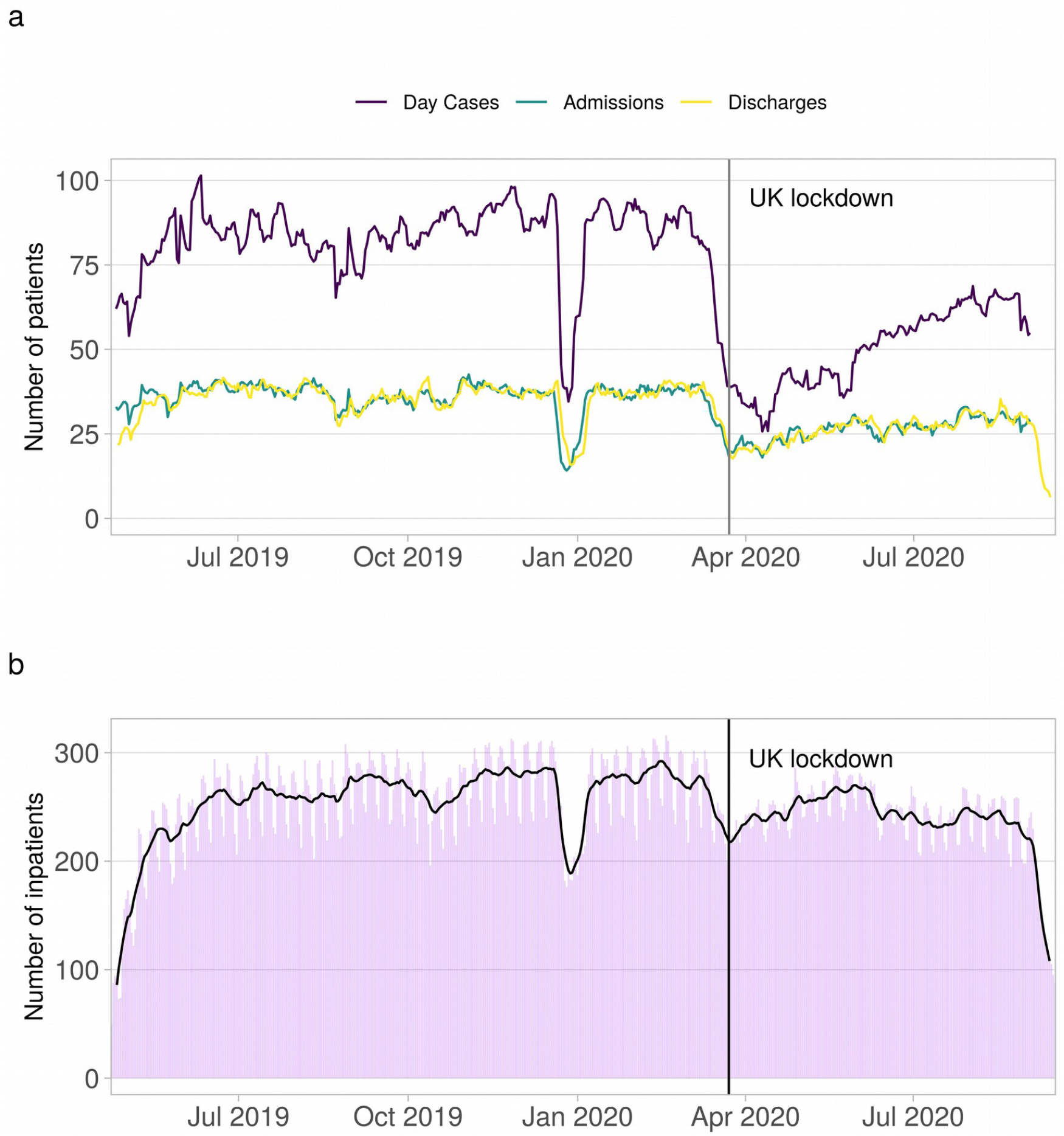
(a) Day cases, inpatient admissions and discharges over time. (b) Inpatient occupancy over time

Patients admitted as day cases during the COVID-19 pandemic were slightly younger (median 7.9 vs 7.0 years), a greater proportion had at least one drug administered (57% vs 68%) and a lower proportion had at least one surgical encounter (15% vs 7.7%) (Table 2).

**Table 2:** A comparison of the characteristics of patients admitted before and after the start of the COVID-19 emergency

|  | Day Case |  |  | Inpatient |  |  |
| --- | --- | --- | --- | --- | --- | --- |
|  | Pre-COVID-19,<br>N =<br>27,348<sup>1</sup> | COVID-19,<br>N = 8,469<sup>1</sup> | p-value<sup>2</sup> | Pre-COVID-19,<br>N =<br>12,008<sup>1</sup> | COVID-19,<br>N = 4,431<sup>1</sup> | p-value<sup>2</sup> |
| Age at admission, years | 7.9 (3.5, 12.6) | 7.0 (3.1, 11.9) | <0.001 | 5.4 (1.8, 10.7) | 5.0 (1.2, 11.2) | 0.006 |
| Male | 14,126 (52%) | 4,306 (51%) | 0.2 | 6,604 (55%) | 2,488 (56%) | 0.2 |
| Length of stay, days | 27,348 (100%) | 8,469 (100%) |  | 3 (2, 6) | 4 (2, 8) | <0.001 |
| Admission type |  |  | <0.001 |  |  | <0.001 |
| Elective - Booked | 2,161 (7.9%) | 350 (4.1%) |  | 1,205 (10%) | 393 (8.9%) |  |
| Elective - Planned | 19,885 (73%) | 6,274 (74%) |  | 6,584 (55%) | 1,967 (44%) |  |
| Elective - Waiting List | 4,684 (17%) | 1,620 (19%) |  | 2,254 (19%) | 539 (12%) |  |
| Emergency Transfer | 497 (1.8%) | 115 (1.4%) |  | 1,058 (8.8%) | 506 (11%) |  |
| Other/missing | 119 (0.4%) | 110 (1.3%) |  | 907 (7.6%) | 1,026 (23%) |  |
| Admission source |  |  | <0.001 |  |  | <0.001 |
| NHS - Ward for General Patients, Younger Physically Disabled, or A&E | 593 (2.2%) | 208 (2.5%) |  | 1,759 (15%) | 1,197 (27%) |  |
| Temporary Place of Residence | 2,292 (8.4%) | 266 (3.1%) |  | 684 (5.7%) | 126 (2.8%) |  |
|  | Pre-COVID-19,<br>N =<br>27,348<sup>1</sup> | COVID-19,<br>N = 8,469<sup>1</sup> | p-value<sup>2</sup> | Pre-COVID-19,<br>N =<br>12,008<sup>1</sup> | COVID-19,<br>N = 4,431<sup>1</sup> | p-value<sup>2</sup> |
| Usual<br>Place of<br>Residence | 24,389<br>(89%) | 7,957<br>(94%) |  | 9,261<br>(77%) | 2,899<br>(65%) |  |
| Other/<br>missing | 72 (0.3%) | 38 (0.4%) |  | 304 (2.5%) | 209 (4.7%) |  |
| Medication<br>administrations<br>per<br>day in<br>hospital | 1.00 (0.00,<br>4.00) | 1.00 (0.00,<br>4.00) | <0.001 | 6.5 (1.6,<br>10.5) | 7.0 (2.5,<br>11.4) | <0.001 |
| At least<br>one<br>medication<br>administration | 15,536<br>(57%) | 5,789<br>(68%) | <0.001 | 9,750<br>(81%) | 3,839<br>(87%) | <0.001 |
| At least<br>one<br>surgical<br>encounter | 3,979<br>(15%) | 651 (7.7%) | <0.001 | 5,698<br>(47%) | 1,664<br>(38%) | <0.001 |

<sup>1</sup>Statistics presented: median (IQR); n (%)
<sup>2</sup>Statistical tests performed: Wilcoxon rank-sum test; chi-square test of independence

There was no statistically significant difference in median age between inpatients admitted before and during the pandemic period. A greater proportion of inpatients were admitted from an NHS ward for general patients, younger physically disabled, or A&E during the pandemic (15% vs 27%) and a lower proportion were admitted from their usual place of residence (77% vs 65%). The median length of stay for inpatients increased by one day from 3 to 4 days. The percentage of inpatients that had at least one medication administration had increased significantly from 81% before the pandemic to 87% during. Simultaneously there was a large drop in the percentage of patients that had at least one surgical encounter during their stay from 47% to 38%.

Most specialities saw a decrease in the number of weekly day cases, but paediatric medical oncology day cases significantly increased (Table 3).

**Table 3:** Change in bed days by treatment speciality

|  | Day Case |  |  | Inpatient |  |  |
| --- | --- | --- | --- | --- | --- | --- |
|  | Pre-COVID-19 , N = 48<sup>1</sup> | COVID-19 , N = 24<sup>1</sup> | p-value<sup>2</sup> | Pre-COVID-19 , N = 48<sup>1</sup> | COVID-19 , N = 26<sup>1</sup> | p-value<sup>2</sup> |
| Blood And Marrow Transplantation | 9 (6, 11) | 7 (4, 8) | 0.006 | 86 (78, 94) | 107 (90, 120) | <0.001 |
| Paediatric Cardiology | 28 (25, 32) | 10 (3, 17) | <0.001 | 63 (52, 76) | 78 (73, 89) | <0.001 |
| Paediatric Clinical Haematology | 50 (44, 55) | 42 (36, 48) | 0.008 | 103 (79, 119) | 104 (85, 129) | 0.4 |
| Paediatric Dermatology | 40 (35, 50) | 14 (12, 29) | <0.001 | 8 (4, 12) | 2 (0, 5) | <0.001 |
| Paediatric Ear Nose And Throat | 13 (10, 16) | 5 (3, 7) | <0.001 | 61 (52, 73) | 26 (17, 36) | <0.001 |
| Paediatric Endocrinology | 15 (13, 17) | 11 (8, 16) | 0.055 | 51 (42, 60) | 19 (5, 25) | <0.001 |
| Paediatric Gastroenterology | 20 (17, 23) | 16 (12, 18) | <0.001 | 73 (59, 82) | 54 (44, 61) | <0.001 |
| Paediatric Intensive Care | 9 (7, 14) | 5 (3, 6) | <0.001 | 308 (283, 382) | 383 (353, 454) | 0.001 |
| Paediatric Medical Oncology | 48 (40, 52) | 54 (50, 60) | <0.001 | 123 (105, 132) | 139 (118, 180) | 0.003 |
| Paediatric Metabolic Disease | 15 (13, 18) | 15 (14, 20) | 0.2 | 26 (18, 31) | 12 (8, 15) | <0.001 |
| Paediatric Nephrology | 48 (44, 52) | 48 (45, 49) | 0.9 | 70 (60, 82) | 58 (48, 65) | <0.001 |
| Paediatric Neurology | 42 (38, 47) | 26 (19, 32) | <0.001 | 42 (31, 49) | 40 (32, 43) | 0.2 |
| Paediatric Neurosurgery | 8 (5, 10) | 8 (4, 11) | 0.8 | 90 (82, 107) | 80 (69, 92) | 0.004 |
|  | Pre-COVID-19 , N =<br>48<sup>1</sup> | COVID-19 , N =<br>24<sup>1</sup> | p-value<sup>2</sup> | Pre-COVID-19 , N =<br>48<sup>1</sup> | COVID-19 , N =<br>26<sup>1</sup> | p-value<sup>2</sup> |
| Paediatric Plastic Surgery | 15 (11, 18) | 4 (2, 8) | <0.001 | 58 (42, 68) | 14 (5, 30) | <0.001 |
| Paediatric Respiratory Medicine | 5 (4, 7) | 1 (1, 2) | <0.001 | 124 (116, 132) | 62 (54, 96) | <0.001 |
| Paediatric Rheumatology | 65 (47, 76) | 24 (19, 28) | <0.001 | 27 (20, 33) | 16 (12, 20) | <0.001 |
| Paediatric Surgery | 13 (8, 15) | 6 (4, 7) | <0.001 | 98 (86, 114) | 79 (60, 94) | <0.001 |
| Paediatric Trauma And Orthopaedics | 17 (14, 19) | 2 (2, 7) | <0.001 | 75 (65, 86) | 18 (13, 26) | <0.001 |
| Paediatric Urology | 57 (47, 62) | 11 (5, 20) | <0.001 | 71 (58, 78) | 40 (28, 55) | <0.001 |
| Other/missing | 60 (52, 69) | 32 (22, 44) | <0.001 | 226 (207, 251) | 282 (263, 333) | <0.001 |

<sup>1</sup>Statistics presented: median (IQR)
<sup>2</sup>Statistical tests performed: Wilcoxon rank-sum test

There was more variation in bed days between specialities for inpatients. Some specialities such as blood and bone marrow transplant, paediatric intensive care and paediatric medical oncology saw large increases in weekly bed days whereas others such as paediatric trauma and orthopaedics, and paediatric respiratory medicine, reported large decreases.

There has been a large reduction in administrations of anaesthesia drugs per 1000 patient bed days for both day cases and inpatients. There was a large reduction in administrations of infection (antimicrobial) drugs per 1000 patient bed days for inpatients (Table 4).

**Table 4:** Medication administrations per 1000 patient days by therapeutic class before and after the start of the COVID-19 emergency

|  | Day Case |  |  | Inpatient |  |  |
| --- | --- | --- | --- | --- | --- | --- |
|  | Pre-COVID-19, N = 333<sup>1</sup> | COVID-19 period, N = 178<sup>1</sup> | p-value<sup>2</sup> | Pre-COVID-19, N = 335<sup>1</sup> | COVID-19 period, N = 178<sup>1</sup> | p-value<sup>2</sup> |
| Anaesthesia | 35.8 (23.5, 48.9) | 24.1 (0.0, 38.8) | <0.001 | 39.2 (21.8, 46.7) | 32.4 (22.5, 40.3) | <0.001 |
| Cardiovascular | 20.0 (13.9, 28.3) | 29.7 (8.8, 44.5) | 0.079 | 152.1 (141.7, 163.4) | 164.3 (152.9, 173.4) | <0.001 |
| Central nervous system | 62.1 (45.2, 79.5) | 47.1 (25.8, 70.5) | <0.001 | 312.8 (300.4, 326.3) | 315.3 (297.2, 327.0) | 0.6 |
| Ear nose and oropharynx | 0.0 (0.0, 0.7) | 0.0 (0.0, 0.0) | <0.001 | 10.2 (8.3, 12.0) | 9.8 (7.5, 12.7) | 0.6 |
| Endocrine | 6.5 (4.1, 9.7) | 7.7 (4.0, 10.1) | 0.5 | 40.6 (37.3, 44.9) | 50.4 (46.2, 54.6) | <0.001 |
| Eye | 0.0 (0.0, 3.1) | 0.0 (0.0, 0.0) | <0.001 | 29.8 (24.5, 36.6) | 31.6 (26.5, 39.5) | 0.003 |
| Gastrointestinal | 1.6 (0.0, 2.9) | 2.1 (0.0, 4.2) | 0.061 | 144.6 (121.5, 157.1) | 129.2 (121.0, 138.9) | <0.001 |
| Immunological products and vaccines | 0.0 (0.0, 0.9) | 1.0 (0.0, 2.3) | <0.001 | 0.4 (0.0, 1.0) | 0.8 (0.1, 1.5) | <0.001 |
| Infections | 5.9 (3.5, 9.5) | 4.7 (0.0, 7.8) | <0.001 | 212.9 (198.4, 227.0) | 235.4 (223.1, 245.7) | <0.001 |
| Malignancy and immunosuppression | 10.9 (6.7, 14.9) | 15.1 (8.9, 23.0) | <0.001 | 24.1 (21.7, 27.4) | 34.7 (30.7, 38.3) | <0.001 |
| Miscellaneous aids materials for diagnostic procedures | 4.1 (0.7, 6.2) | 4.8 (0.0, 7.1) | 0.3 | 2.0 (0.8, 2.8) | 1.8 (0.9, 2.8) | >0.9 |
|  | Pre-COVID-19,<br>N = 333<sup>1</sup> | COVID-19<br>period, N = 178<sup>1</sup> | p-value<sup>2</sup> | Pre-COVID-19,<br>N = 335<sup>1</sup> | COVID-19<br>period, N = 178<sup>1</sup> | p-value<sup>2</sup> |
| Miscellaneous appliances | 0.0 (0.0, 0.0) | 0.0 (0.0, 0.0) | 0.068 | 3.0 (1.8, 4.0) | 4.2 (3.2, 5.1) | <0.001 |
| Musculoskeletal and joint diseases | 13.7 (6.1, 18.8) | 6.2 (0.0, 10.4) | <0.001 | 34.9 (31.6, 38.5) | 29.1 (25.8, 32.3) | <0.001 |
| Nutrition and blood | 41.3 (32.7, 59.2) | 61.5 (28.8, 83.6) | 0.11 | 218.8 (206.7, 232.3) | 234.8 (223.3, 254.0) | <0.001 |
| Obstetrics gynaecology and the urinary tract | 0.0 (0.0, 0.7) | 0.0 (0.0, 1.1) | 0.048 | 5.0 (3.8, 6.5) | 2.6 (1.7, 3.7) | <0.001 |
| Parenteral nutrition | 0.0 (0.0, 0.0) | 0.0 (0.0, 0.0) |  | 23.7 (20.6, 26.3) | 22.4 (20.7, 25.0) | 0.007 |
| Probiotics | 0.0 (0.0, 0.0) | 0.0 (0.0, 0.0) | 0.5 | 1.0 (0.7, 1.3) | 1.6 (1.3, 1.8) | <0.001 |
| Respiratory | 4.7 (2.0, 7.1) | 7.0 (0.0, 11.8) | <0.001 | 62.7 (57.6, 69.1) | 59.5 (56.0, 64.6) | <0.001 |
| Skin | 0.0 (0.0, 0.8) | 0.0 (0.0, 0.0) | <0.001 | 24.1 (21.1, 27.6) | 21.7 (18.3, 24.7) | <0.001 |
| Stoma appliances | 0.0 (0.0, 0.0) | 0.0 (0.0, 0.0) | 0.2 | 2.1 (1.3, 3.1) | 1.2 (0.4, 2.1) | <0.001 |
| Other missing | 43.1 (32.7, 50.0) | 32.4 (23.0, 40.0) | <0.001 | 22.0 (19.5, 24.7) | 19.5 (17.2, 22.7) | <0.001 |

<sup>1</sup>Statistics presented: median (IQR)
<sup>2</sup>Statistical tests performed: Wilcoxon rank-sum test

## Discussion

The COVID-19 pandemic has caused major disruptions to health care delivery in the UK. This study describes hospital level activity, patient characteristics and prescribing patterns for patients admitted to a specialist children’s hospital before and during the COVID-19 pandemic. Understanding how care changed in the early stages of the pandemic will be important to prepare for future emergencies.

The findings of this study show that there were significant reductions in hospital activity at a specialist children’s hospital during the COVID-19 pandemic compared to the baseline. In particular, day case activity dropped by around 37% per week (from 588 before to 370). Both admissions and discharges for inpatients decreased, leading to a small overall reduction in hospital bed days but no reduction in hospital bed nights.

Overall weekly medication administrations and surgical encounters decreased for both day cases and inpatients but when scaled by bed days, weekly medication administrations per bed day increased by 6%, which likely reflects changes to the patient population, with only those children with severe conditions undertaking treatments during the pandemic.

At a patient level, more inpatients were given at least one drug during their stay during the pandemic (81% vs 87%) and a lower percentage had at least one surgical encounter during their stay (47% vs 38%).

Other studies have reported decreases and delays in A&E attendances for children and young people ([6], [7], [13]) both in the UK and abroad as a consequence of the COVID-19 emergency. Common explanations include potential lower morbidity during the pandemic and an avoidance of hospital visits either to avoid being exposed to COVID-19 or to avoid burdening the NHS. Care utilisation at specialist hospitals like GOSH is likely to be driven by different factors, given the highly specialist nature of the care and the absence of an A&E department.

A number of factors will have contributed to the variation in changes to patient bed days for different specialities, with different implications. For some specialities the decrease will be due to elective care being postponed as a result of the pandemic. However, such patients will continue to require treatment in the near future, an increase in demand that will lead to different challenges. The large increases in intensive care patients during the COVID-19 pandemic period are likely the result of transfers from other hospitals, as is the increase in number of cancer patients. For other specialities such as paediatric trauma and orthopaedics and paediatric respiratory medicine, the reduction in bed days is likely a consequence of a decrease in demand due to behavioural changes during lockdown. One study reported reduction in paediatric admissions for acute asthma at a London teaching hospital, with suggested reasons including distancing from peers leading to reduced transmissions of respiratory viruses [14]. There are also reports of reduced trauma surgeries as road traffic accidents and sporting injuries were less prevalent during the pandemic [15].

Drug administration data provides information regarding how medication treatment changed during the pandemic. Some changes reflect alterations in case mix, such as the large reduction in anesthesia use in association with reduced surgical encounters and the increase in malignancy and immunosuppression drugs as a result of the increase in cancer patients referred to the specialist centre during COVID-19. The increase in antimicrobial drugs is more difficult to interpret since this may represent several factors such as changing referral patterns or increased antimicrobial prophylaxis and use in patients with more complex healthcare requirements such as in oncology. Pharmacist working patterns changed during the pandemic which could have influenced antimicrobial stewardship activities. There are concerns that the COVID-19 pandemic will increase levels of antimicrobial resistance in part due to relaxing of antimicrobial stewardship activities[16].

A major strength of this study is the use of a large routinely collected, comprehensive patient-level data set with very recent data included. Hospital medication data is an underused resource in health care research [17]. The use of medication administration data in this study allows us to go beyond hospital use and quantify changes to medical treatments during the pandemic. Our study population is representative of a specialist children’s hospital, and the longer time period covered allows us to understand the effect of COVID-19 not just during the height of the pandemic but also the prolonged effect on health care.

One limitation is that the pre-COVID-19 period does not cover the identical calendar months, making it impossible to adjust for factors related to seasonality. Similarly, an extended time period would allow more robust statistical timeseries analyses. As a sensitivity analysis, we used historical admission data from 2016 to 2018 and found similar trends for hospital admissions and discharges. Though this was not possible for all metrics in this study due to differences between the old and new data systems, this sensitivity analysis indicates that the pre-pandemic period is representative of baseline activity. It should also be highlighted that the data we present here is from a highly specialist children’s hospital and patients are not representative of the general paediatric population or children in primary or secondary care settings. We were unable to say whether the patterns of utilisation were appropriate clinically for the needs of the patients or assess the experience of the children and their carers.

The COVID-19 pandemic has caused an unprecedented impact on health care systems in the UK and globally. The findings of the current study confirm a significant decrease in the number of day cases being treated at a specialist children’s hospital but inpatient activity for children with rare and complex disorders was less affected. There is however wide variation between treatment specialities, leading to some patient groups having been virtually unaffected and others severely affected. Whilst overall prescribing activity reduced, the average number of medications per inpatient bed day increased. These data confirm the complex pattern of implications on specialist services and should be useful for planning for the impact of future emergencies and resolution strategies.

### What is already known on this topic?

- There are growing concerns that children are negatively affected by changes to society despite not in general being affected by the disease itself
- There was a reduction in paediatric emergency attendances during the COVID-19 pandemic.

### What this study adds?

- Our analysis shows that the COVID-19 pandemic affected care at a specialist children’s hospital, with reduced surgical activity the most marked effect.
- Per-patient medication use rose, including increased use of antimicrobials.

## Data Availability

Data may be obtained from a third party and are not publicly available.

## Code

https://github.com/emmavestesson/COVID-prescribing

